# ANO1 expression is associated with male survival in lung squamous cell carcinoma

**DOI:** 10.1101/2024.08.14.24311973

**Authors:** Oluwadamilare I. Afolabi

## Abstract

Lung cancer is the leading cause of cancer deaths, with lung squamous cell carcinoma (LUSC) accounting for a substantial proportion of cases. LUSC exhibits significant variability in patient outcomes, influenced by clinicopathological factors such as stage, age, and sex, with men exhibiting higher rates of incidence and poorer outcomes compared to women. Therefore, prognosis modeling and customized approaches to LUSC therapy require the characterization of the biological mechanisms that differentiate tumors and patient outcomes across sexes. Using data from The Cancer Genome Atlas (TCGA), this study characterized gene expression patterns that distinguish male and female LUSC. Specifically, differential expression, survival, and Cox regression analyses assessed the prognostic value of ANO1 (Anoctamin 1) expression and methylation in LUSC. Analyses uncovered a significant overexpression of ANO1 in a subset of male LUSC tumors compared to a much lower expression in normal lung and female LUSC tumors. High ANO1 expression was associated with poor survival outcomes in male subjects. High ANO1 gene body methylation mirrored gene expression and was similarly associated with poor survival outcomes. ANO1’s prognostic value remained significant in a multivariate Cox regression analysis, establishing it as an independent prognostic biomarker. ANO1’s marked sex-specific differences in expression and prognostic value indicate its role in the sex disparity of LUSC survival, highlighting its potential as a biomarker and a target for sex-specific customized therapies.

## Introduction

Lung cancer remains the leading cause of cancer-related mortality globally, with lung squamous cell carcinoma (LUSC) constituting a substantial proportion of cases [1]. Despite advancements in treatment, the expected 5-year survival rate for LUSC is about 22%, with improved outcomes for those diagnosed at earlier stages, younger ages, and for women compared to men [2, 3]. While variations in risk factors account for some of this sex disparity, the survival difference persists even after adjusting for risk exposure. Consequently, sex-specific studies on LUSC biology, disease progression, and survival are required. Evidence from other cancer types implicates various gene expressions and mutations in sex-based differences [4, 5]. However, the molecular mechanisms underlying the disparities in LUSC incidence and prognosis remain poorly understood.

In normal cells, proto-oncogenes regulate cell growth, differentiation, and other functions. When overexpressed, oncogenes can lead to uncontrolled cell proliferation and other hallmarks central to tumorigenesis and cancer progression. Numerous oncogenes have been identified as prognostic biomarkers and therapeutic targets in various cancers [6, 7, 8]. However, the heterogeneity of LUSC complicates the identification of reliable predictors of clinical outcomes [9, 10]. Moreover, the genetic variations between sexes that contribute to differences in survival rates are even less characterized. Further research is needed to elucidate these molecular differences, as they can help improve personalized treatment strategies and ultimately, prognosis for LUSC [11].

This study leveraged high-throughput gene expression data from The Cancer Genome Atlas (TCGA) to evaluate sex-biased differential gene expression between LUSC and matched adjacent-normal lung tissues. Specifically, the study focused on ANO1 (Anoctamin 1) due to its consistently low expression in adjacent-normal male and female tissues, its differential expression in male versus female LUSC tissues, and the significant variation of its expression within male LUSC tissues. The study demonstrated the value of ANO1 expression and methylation for predicting survival in male LUSC subjects, suggesting its role in the sex disparity of LUSC survival and consistent with its established involvement in other cancers.

## Materials and Methods

### Data acquisition and preprocessing

RNA-seq and DNA methylation array data for 341 subjects diagnosed with primary Stage I and II lung squamous cell carcinoma (dbGaP accession phs000178.v11.p8) were retrieved from TCGA Genomic Data Commons [12]. The cohort comprised 248 male and 93 female subjects. Matched adjacent-normal lung samples were also obtained for a subset of subjects, including 10 females and 29 males (Fig. S1).

For differential gene expression (DGE) analyses, matched sets of 10 male and 10 female LUSC samples were selected alongside their corresponding adjacent-normal lung tissues. Sample matching was based on clinical characteristics and risk exposure, with the absence of confounding factors confirmed using T-tests for continuous variables and Chi-squared tests for discrete variables (Fig. S2).

### Differential gene expression analyses

All DGE analyses were conducted using DESeq2 [13]. First, genes with very low counts, specifically those with fewer than 10 counts in all samples of the smallest sample group, were filtered out. To detect genes dysregulated in LUSC, differentially expressed genes (DEGs) were first identified between tumor and matched adjacent-normal samples for the combined male and female cohort. Subsequently, sex-specific dysregulation was explored by identifying DEGs between male and female tumor samples. Finally, to investigate ANO1-associated dysregulation, male subjects were stratified into low and high ANO1 expression groups based on an optimal threshold identified using survminer [14]. DEGs were identified between these high and low ANO1 expression groups. An absolute fold change of four and an adjusted p-value of 0.05 threshold were used to select significant DEGs. Volcano plots were utilized to visualize the expression patterns of these DEGs. Wilcoxon signed-rank tests were employed to compare the distributions of DEGs across various conditions.

### Functional enrichment analysis

Gene Ontology (GO) functional enrichment analysis was performed using clusterProfiler [15] to identify biological processes associated with LUSC biology. A q-value threshold of 0.01 was applied to select significantly overrepresented GO processes in sex-shared and sex-specific LUSC-dysregulated genes. Sex-specific DEGs were annotated using biomaRt [16] to distinguish between autosomal and sex chromosome genes, ensuring that sex-linked genes were appropriately accounted for, thereby providing a clearer understanding of differences due to LUSC and not sex alone.

### Methylation analysis

DNA methylation patterns in the ANO1 locus were investigated, focusing on CpGs in the TSS1500 (distal promoter; 200-1,500 bp upstream of the transcriptional start site), TSS200 (proximal promoter; 0-200 bp upstream of TSS), Exon 1 (first exon; between the start codon and the end of the first exon), and Body (gene body; between the end of the first exon and the stop codon) [17]. Pearson correlation analysis between methylation and gene expression was performed for the proximal promoter and gene body regions. A Methylation Index (MI) was calculated, accounting for both gene body and proximal promoter methylation in each sample. Subjects were stratified by proximal promoter methylation, gene body methylation, and MI using optimal thresholds identified by survminer. Wilcoxon signed-rank tests were employed to compare the distributions of CpG methylation across various conditions.

### Survival outcome analysis

Kaplan-Meier survival analysis was conducted using the survival [18] and survminer packages to evaluate the prognostic significance of ANO1 expression in male subjects. A log-rank test compared survival distributions between different ANO1 expression groups. Subsequently, survival analyses and log-rank tests were conducted to assess the impact of ANO1 methylation on survival in the same cohort. A multivariate Cox proportional hazards model incorporating clinicopathological features and methylation status was developed and evaluated using a forest map. Additionally, the performance of a univariate Cox model based solely on MI was assessed using the pROC package [19], allowing for a comprehensive evaluation of the predictive power of MI in isolation. These analyses collectively provide a robust framework for understanding the role of ANO1 in male LUSC survival.

## Results

### Sex-specific dysregulation in LUSC

A comprehensive DGE analysis was performed to identify dysregulated genes in LUSC. For the combined cohort of male and female subjects, a total of 2,672 genes were found to be differentially expressed between tumor and matched adjacent-normal samples, with an absolute fold change threshold of 4 and an adjusted p-value threshold of 0.05 (Fig. 1a). The analysis revealed 1,340 upregulated and 1,332 downregulated genes in tumor samples compared to adjacent-normal tissues. Notably, known oncogenes, including FAM83B, KCNMB2-AS1, and TFAP2A, were the most significantly upregulated genes, indicating their potential roles in LUSC pathology. Conversely, known tumor suppressors, MYCT1, LDB2, and NOSTRIN, were the most significant downregulated coding genes, suggesting that their reduced expression may contribute to loss of growth control and tumor progression in LUSC (Supplementary Table S1). Functional enrichment analyses of these DEGs revealed several overrepresented biological processes (Supplementary Table S2). In upregulated DEGs, processes related to cell division, DNA replication, epidermal development, branching morphogenesis, and mitotic cell cycle processes were significantly overrepresented. In contrast, processes linked to vasculature, immune system, and cell-cell adhesion were enriched in downregulated DEGs.

**Fig. 1.**
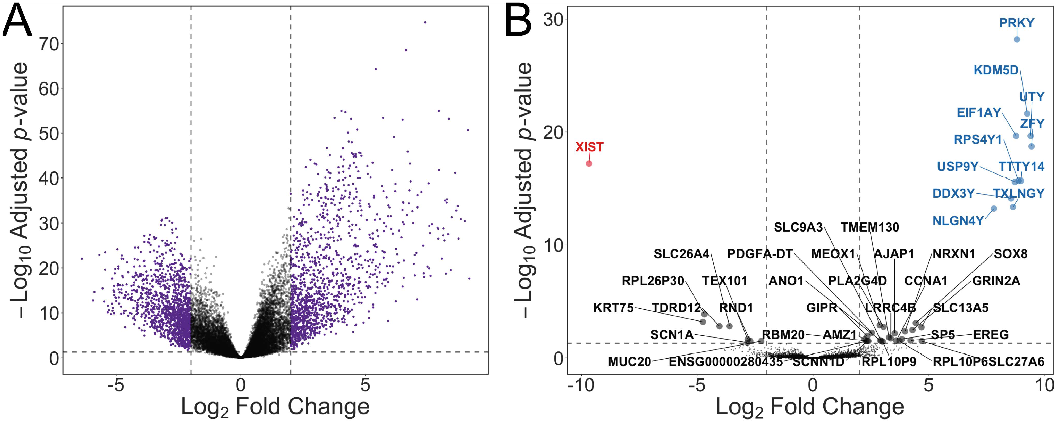
Differential gene expression in LUSC. **(A)** Volcano plot of DEGs in combined male and female tumor versus adjacent-normal tissue samples. DEGs are colored in purple. **(B)** Volcano plot of DEGs in male versus female tumor samples. X-linked DEGs are colored in red, Y-linked in blue, and autosomal in black. All DEGs were selected using an absolute fold change of 4 and an adjusted p-value of 0.05 threshold.

DGE analysis revealed distinct gene expression patterns between male and female LUSC tumors. A total of 43 DEGs were identified, with 34 genes upregulated and 9 genes downregulated in male tumors compared to female tumors (Fig. 1b, Supplementary Table S3). Notably, the X-linked gene XIST was significantly downregulated in male subjects. Several Y-linked genes, including PRKY, KDM5D, and UTY, were upregulated, reflecting expected sex chromosome-specific expression differences. Among autosomal genes, RPL26P30, KRT75, SOX8, and MEOX1 were the most consistently upregulated in male LUSC tissue samples, suggesting their integral roles in LUSC biology.

### Prognostic significance of ANO1 expression

ANO1 was identified as a gene of interest among DEGs because of its distinctive expression pattern. (Fig. S3). Expression analysis revealed that ANO1, while marginally expressed in normal male and female adjacent-normal lung tissue, was significantly upregulated in male compared to female LUSC tumors (Fig. 2a). When stratified by ANO1 expression, a clear distinction exists between adjacent-normal tissue, tumors with low ANO1 expression, and tumors with high ANO1 expression, suggesting the gene’s potential role in tumor biology (Fig. 2b). High ANO1 expression was observed to correlate with poorer overall survival in male subjects (log-rank p-value = 0.04), suggesting its potential as a prognostic biomarker for male LUSC (Fig. 2c). ANO1, a promoter of cell proliferation, has previously been implicated in tumor progression and metastasis in various cancers [20].

**Fig. 2.**
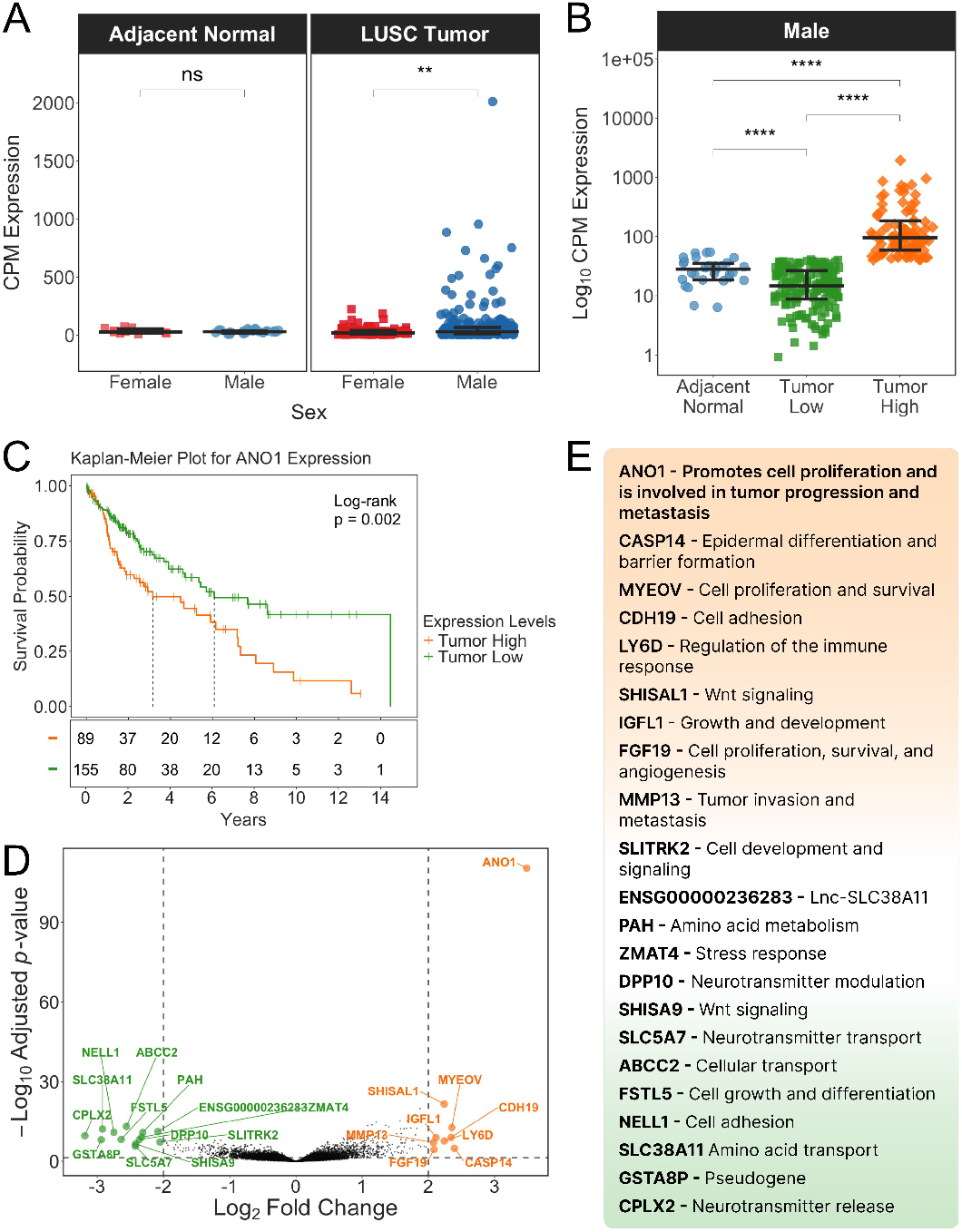
Differential expression and prognostic significance of ANO1 in LUSC. **(A)** Dot plots comparing CPM-normalized ANO1 expression in adjacent-normal tissue and tumor samples of male and female subjects. **(B)** Dot plots comparing log _10_ CPM-normalized ANO1 expression in adjacent-normal tissue, low, and high ANO1-expressing tumor samples of male subjects. Dots, horizontal lines, and error bars represent individual subjects, group medians, and group IQRs, respectively. **(C)** Kaplan-Meier survival plot comparing overall survival between male subjects with high and low ANO1-expressing tumors. Vertical dotted lines represent median survivals. **(D)** Volcano plot of DEGs in high versus low ANO1-expressing male tumor samples. Upregulated DEGs are colored in orange and downregulated in green. DEGs were selected using an absolute fold change of 4 and an adjusted p-value of 0.05 threshold. **(E)** DEGs in high versus low ANO1-expression groups sorted by fold change.

To further explore the implications of ANO1 expression in LUSC, a differential expression analysis was conducted between tumors with high and low ANO1 expression. A total of 22 DEGs were identified, with 9 genes upregulated and 13 genes downregulated (Fig. 2d, Supplementary Table S4). Functional annotation of the DEGs revealed a substantial involvement of upregulated genes in key cancer-related biological processes (Fig. 2e). Other upregulated genes such as CASP14, MYEOV, and CDH19 are involved in epithelial differentiation, cell proliferation, and cell adhesion, respectively. Downregulated genes including SLC38A11, NELL1, and CPLX2 are linked to amino acid transport, cell adhesion, and neurotransmitter release, indicating their role in tumor suppression or an alternative pathway that may contribute to a less aggressive phenotype.

### ANO1 differentially methylated with gene expression

Significant differences in methylation of the ANO1 locus were observed between high and low ANO1 expression groups (Fig. S4). In the proximal promoter region, a slight yet significant reduction in methylation levels was observed in the high expression group versus the low expression group, despite the generally low methylation of this region (Fig. 3a). A more detailed analysis of individual CpGs within the region revealed that while some sites are differentially methylated, others are not (Fig. 3b). In contrast, ANO1 gene body demonstrated marked and significantly higher methylation in the high expression group compared to the low expression group, coupled with overall higher methylation levels. Individual CpGs in the gene body revealed a consistent pattern of increased methylation across the fourteen most differentially methylated CpGs within the gene body (Fig. 3c). This pronounced increase in methylation within the gene body and marginal decrease in the proximal promoter region suggests that ANO1 locus methylation is chiefly a consequence of elevated transcription. The distal promoter and first exon regions did not exhibit significant differences in methylation between the high and low expression groups (Fig. 3a, Fig. S4).

**Fig. 3.**
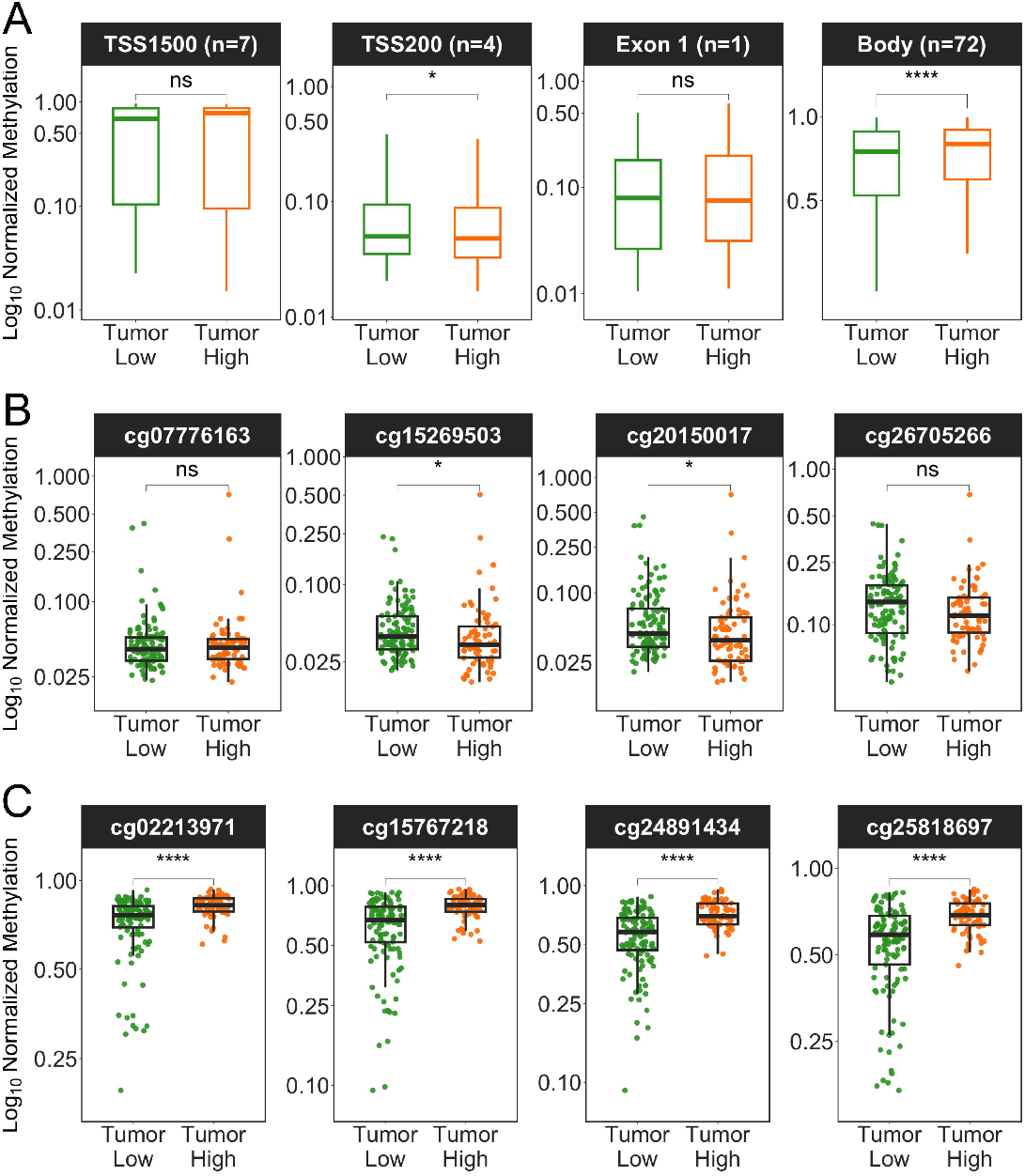
ANO1 differentially methylated with gene expression. **(A)** Boxplots comparing log_10_ normalized methylation of distal promoter (TSS1500), proximal promoter (TSS200), first exon (Exon 1), and gene body (Body) regions of high and low ANO1-expressing male tumor samples. **(B)** Boxplots comparing log_10_ normalized methylation of the non-zero CpGs in the proximal promoter region of high and low ANO1-expressing male tumor samples. **(C)** Boxplots comparing log_10_ normalized methylation of the top 4 most differentially methylated CpGs in the gene body region of high and low ANO1-expressing male tumor samples. Dots represent individual subjects.

### Prognostic significance of ANO1 methylation

A Pearson correlation analysis revealed a strong positive correlation (R = 0.515, p-value < 0.001) between gene body methylation and gene expression (Fig. 4a). Conversely, a weak negative correlation (R = -0.097, p-value < 0.185) was observed between proximal promoter methylation and gene expression. These results confirm earlier observations of methylation differences between high and low ANO1 expression groups (Fig. 3a). Next, tumor samples were stratified by proximal promoter methylation, gene body methylation, or MI (Fig. 4b). Compared with the proximal promoter (log-rank p-value = 0.2) and gene body (log-rank p-value = 0.097) methylation, MI performed best (log-rank p-value = 0.0053) at predicting survival, with high MI correlating with poorer overall survival (Fig. 4c). Together, these results suggest that ANO1 methylation mirrors gene expression and is potentially as effective a biomarker as ANO1 expression.

**Fig. 4.**
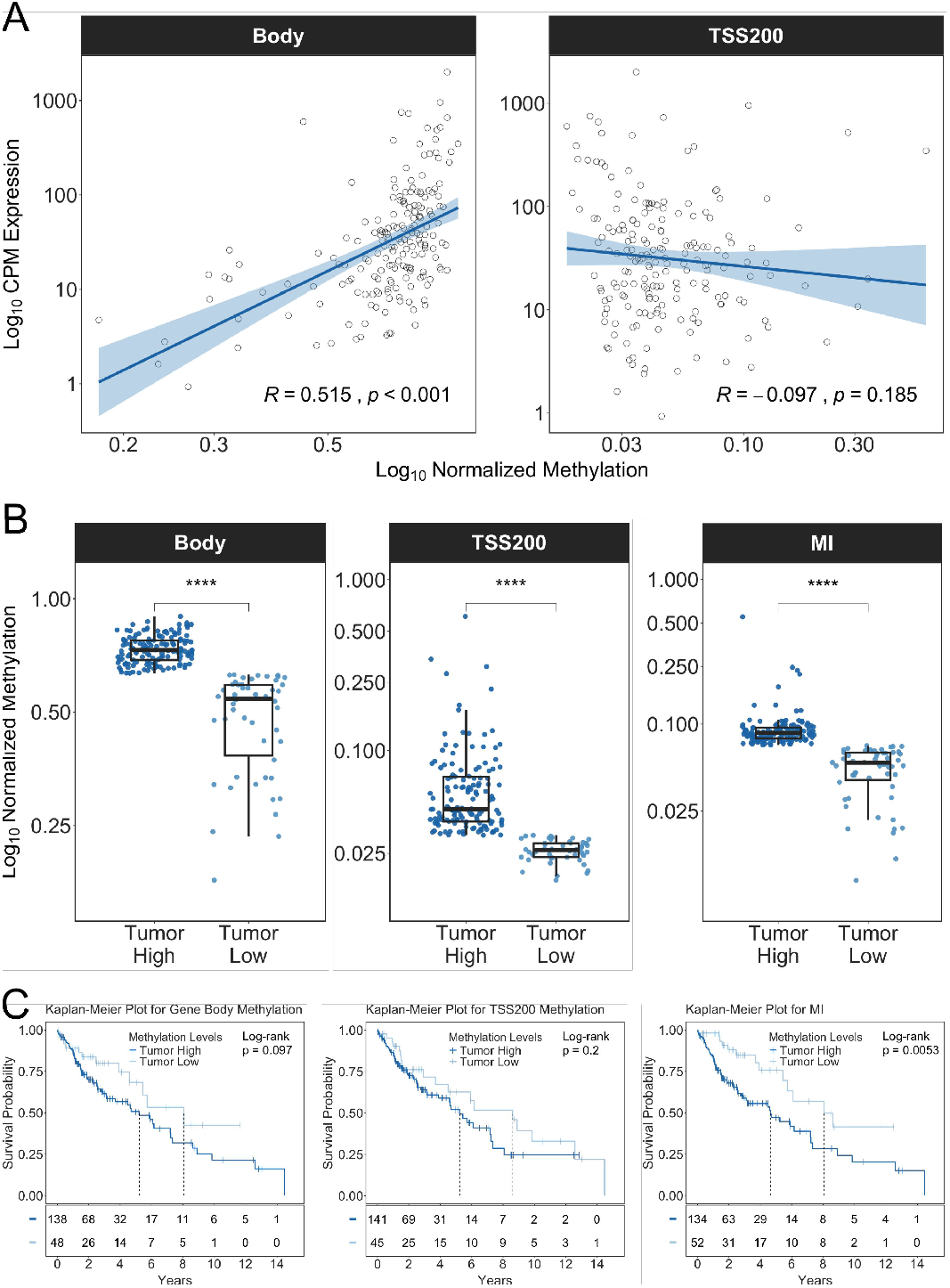
Prognostic significance of ANO1 methylation in LUSC. **(A)** Correlation plots of log_10_ CPM-normalized gene expression and log_10_ normalized methylation of gene body and proximal promoter regions of ANO1 in male tumor samples. Dots represent individual CpGs in individual subjects. **(B)** Boxplots illustrating stratification of male subjects by ANO1 gene body methylation, proximal promoter methylation, or methylation index (MI) of tumor samples. Dots represent individual subjects. **(C)** Kaplan-Meier survival plots comparing overall survival in male subjects stratified by ANO1 gene body methylation, proximal promoter region methylation, or MI of tumor samples. Vertical dotted lines represent median survivals.

Using Cox proportional hazards, a multivariate analysis was conducted to assess the correlation between ANO1 methylation status and survival. The hazard ratios for proximal promoter methylation, gene body methylation, and MI were 1.45 (95% CI 0.82-2.57, p-value = 0.207), 0.94 (95% CI 0.41-2.12, p-value = 0.878), and 2.55 (95% CI 1.12-5.78, p-value = 0.025), respectively (Fig. Sa). A univariate model was used to assess the prognostic significance of MI alone. The analysis revealed a significant correlation (risk ratio = 2.34, 95% CI 1.28-4.28, p-value = 0.006) between survival and MI (Fig. 5b). The prognostic value of MI was further confirmed using a time-dependent ROC analysis, demonstrating accurate survival prediction with AUCs of 0.968, 0.935, 0.921, and 0.866 at 1, 3, 5, and 8 years, respectively.

**Fig. 5.**
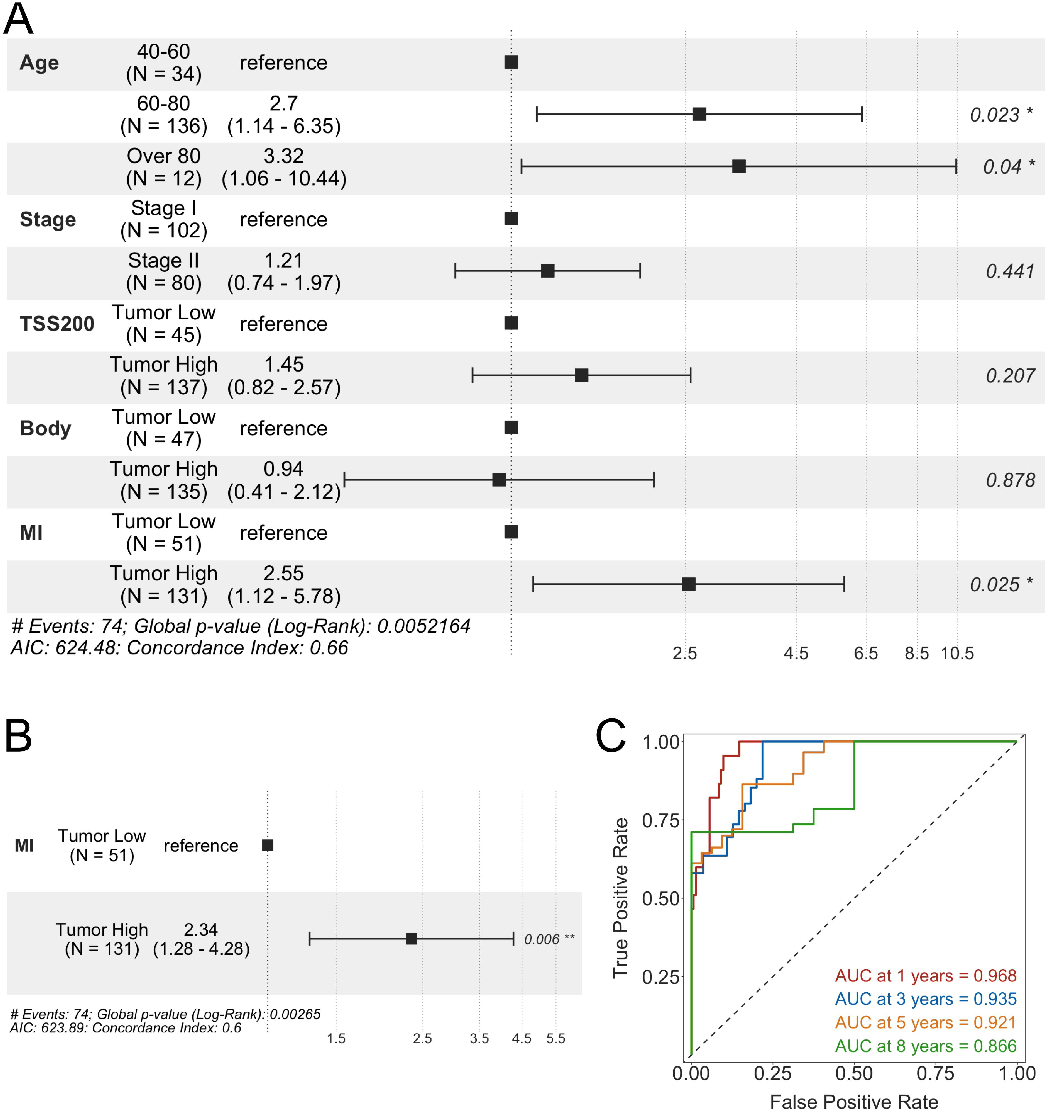
ANO1 methylation is an independent predictor of male LUSC survival. **(A)** Multivariate Cox regression analysis of methylation statuses and clinical variables associated with survival. **(B)** Univariate Cox regression analysis of MI. Dots and error bars represent risk ratios and confidence intervals, respectively. **(C)** Time-dependent ROC curves for MI at 1, 3, 5, and 8 years.

## Discussion

LUSC is characterized by significant variability in patient outcomes, influenced by a range of clinicopathological factors such as stage, age, risk exposure, tumor characteristics, and sex [21] Traditional models based on these factors have been useful but are often insufficient for explaining patient outcomes due to the complex biology of LUSC. This underscores the need for prognostic markers that can provide more accurate predictions and guide customized treatment approaches. Recent advances have highlighted the importance of molecular biomarkers in the prognosis and treatment of various cancers [22, 23, 24]. These biomarkers offer insights into the underlying biology of tumors, aiding personalized treatment.

This investigation examined early primary LUSC tumor datasets from the TCGA database for sex-specific survival genes. Analysis revealed that ANO1 is overexpressed in a subset of male LUSC tumors but only marginally expressed in normal lung tissue. Results also revealed sex-specific differences in ANO1 expression and associated high ANO1 expression and methylation with poor survival outcomes. This association remained significant after adjusting for clinicopathological factors in a multivariate Cox regression analysis, proposing ANO1 as an independent prognostic marker. A methylation index, that accounts for both gene body and proximal promoter methylation, demonstrated accurate survival prediction at 1, 3, 5, and 8 years. These findings implicate ANO1 in sex-biased LUSC survival and highlight its role as a potential biomarker for identifying high-risk LUSC subjects.

In addition to its predictive value, ANO1’s role in tumorigenesis and cancer progression has been well-documented [20, 25, 26]. ANO1 influences various cellular processes including proliferation, migration, and invasion which are critical for cancer progression and metastasis. This study adds to this body of knowledge by demonstrating ANO1’s overexpression as not merely a byproduct of tumorigenesis but a driver of poor clinical outcomes, particularly in male LUSC subjects. Integrating ANO1 into clinical practice as a prognostic biomarker could significantly improve patient management. For instance, high ANO1 expression could identify patients who would benefit from more aggressive treatment regimens or closer surveillance. Moreover, ANO1-targeted therapies could be developed to inhibit its oncogenic functions, thereby improving subject survival [27, 28]. This aligns with the growing trend of precision oncology, where molecular profiling guides treatment decisions to achieve better clinical outcomes [29, 30].

This study further highlights the importance of sex-specific analyses in cancer research. The differential impact of ANO1 expression in male and female subjects underscores the need for sex-conscious research and clinical practice. This is particularly relevant given the known sex bias in LUSC prognosis and the distinct biological mechanisms that may underlie various cancer progressions in men and women [31, 32]. However, the study’s reliance on computational analyses necessitates validation to provide a more comprehensive understanding of ANO1’s role in LUSC.

## Conclusions

Through various analyses, including gene expression analysis, methylation analysis, survival analysis, and machine learning, ANO1 was implicated in sex-biased LUSC survival. ANO1 expression was elevated in a subset of male LUSC tumors, with the increase correlating with unfavorable prognosis. This trend was mirrored by ANO1 methylation, as high ANO1 methylation also correlated with poor survival outcomes. Together, these results suggest that ANO1 holds significant potential as a prognostic biomarker and as a possible target for sex-specific customized LUSC therapy.

## Acknowledgments

The results shown here are wholly based on available data from the TCGA Research Network: https://www.cancer.gov/ccg/research/genome-sequencing/tcga.

## Ethics Approval and Consent

Not applicable.

## Data Availability

All the data used in this study are openly available in the TCGA Genomic Data Commons at https://portal.gdc.cancer.gov, dbGaP accession phs000178.v11.p8.

