## Supplemental Figures S1-S4 for "ANO1 expression is associated with male survival in lung squamous cell carcinoma"

### Supplementary Figures

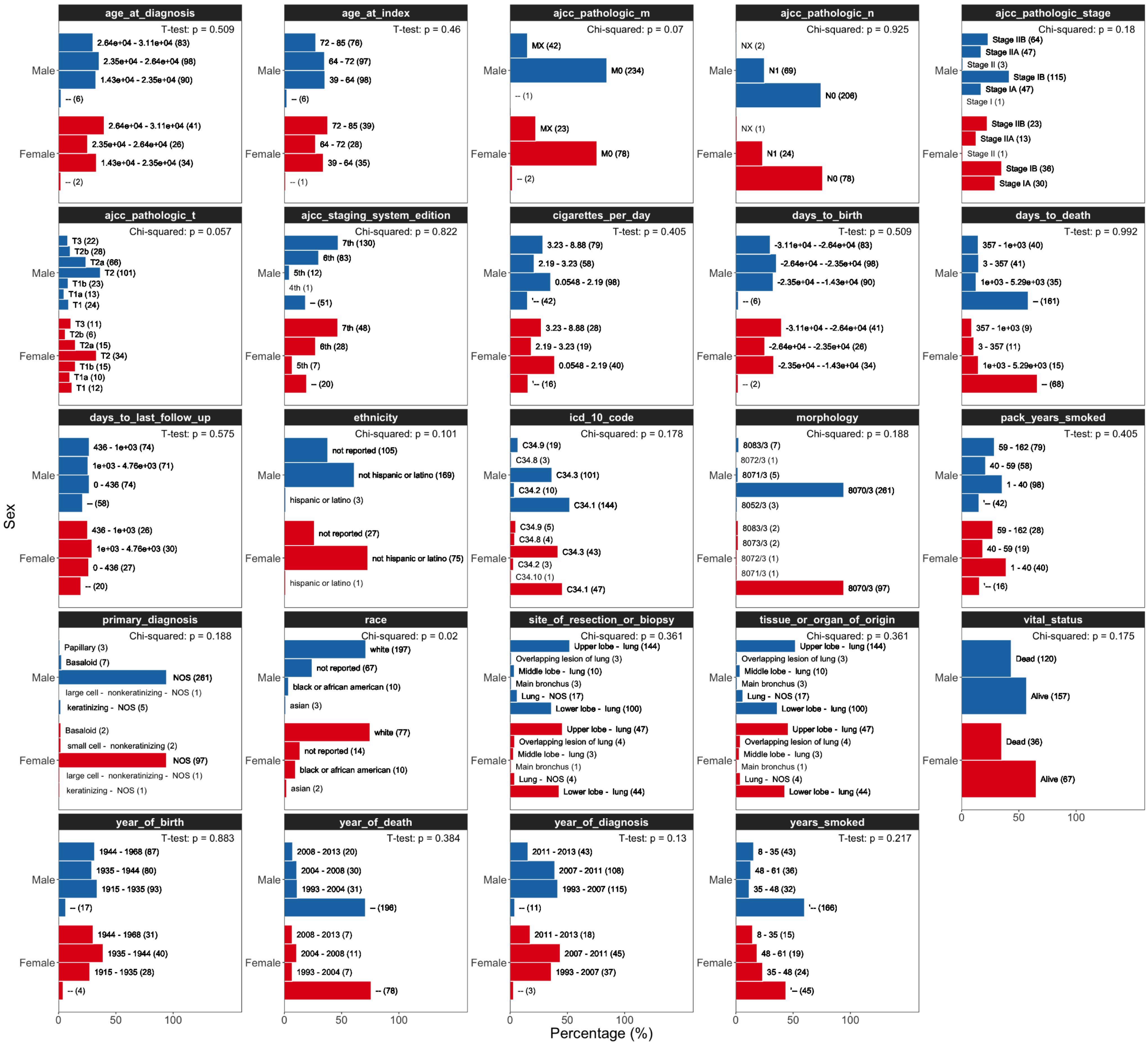


**Fig. S1**. Clinicopathological factors and risk exposures of subjects in the TCGA LUSC cohort.

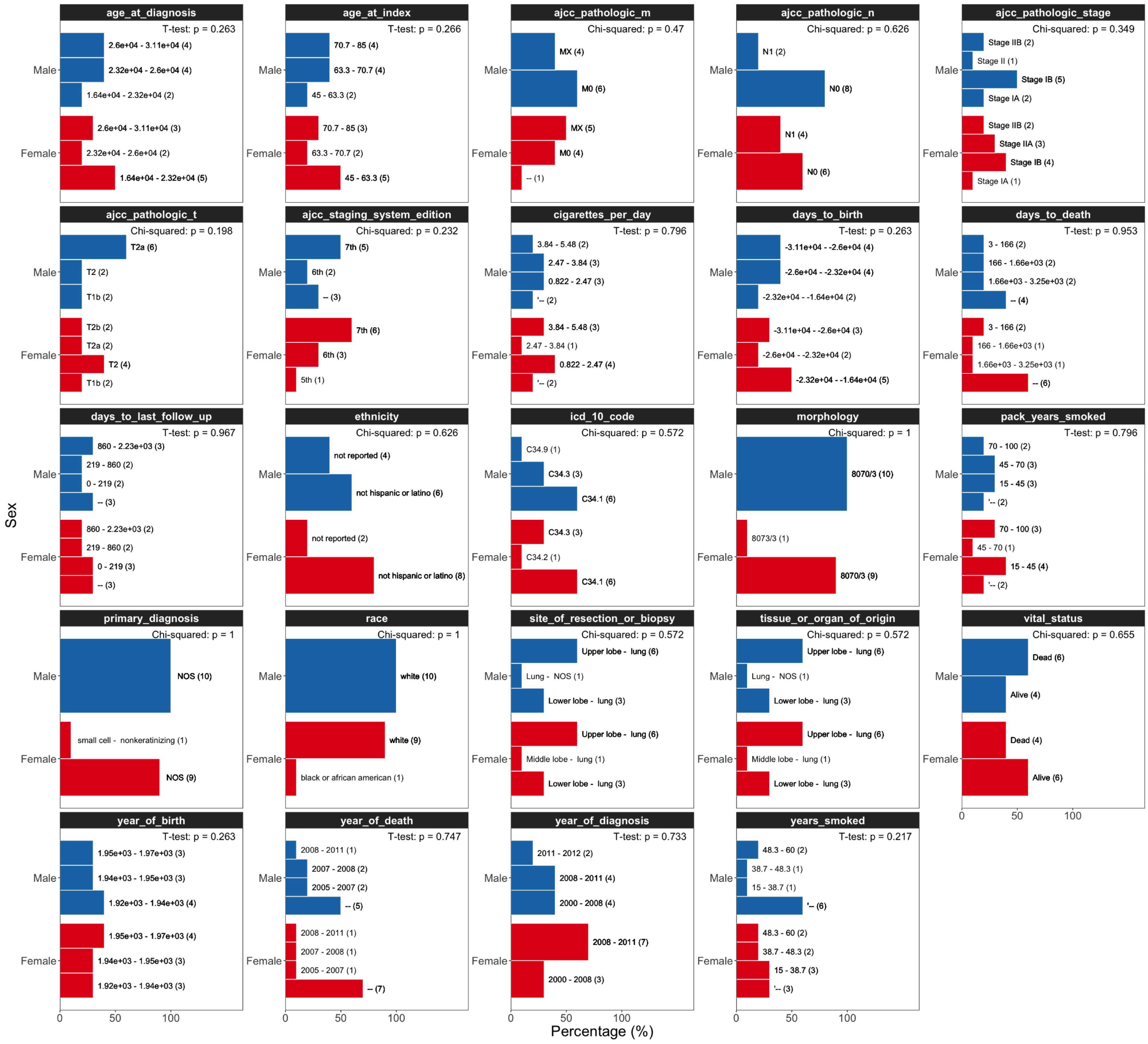


**Fig. S2**. Clinicopathological factors and risk exposures of matched sets of 10 male and 10 female subjects used for DGE analyses.

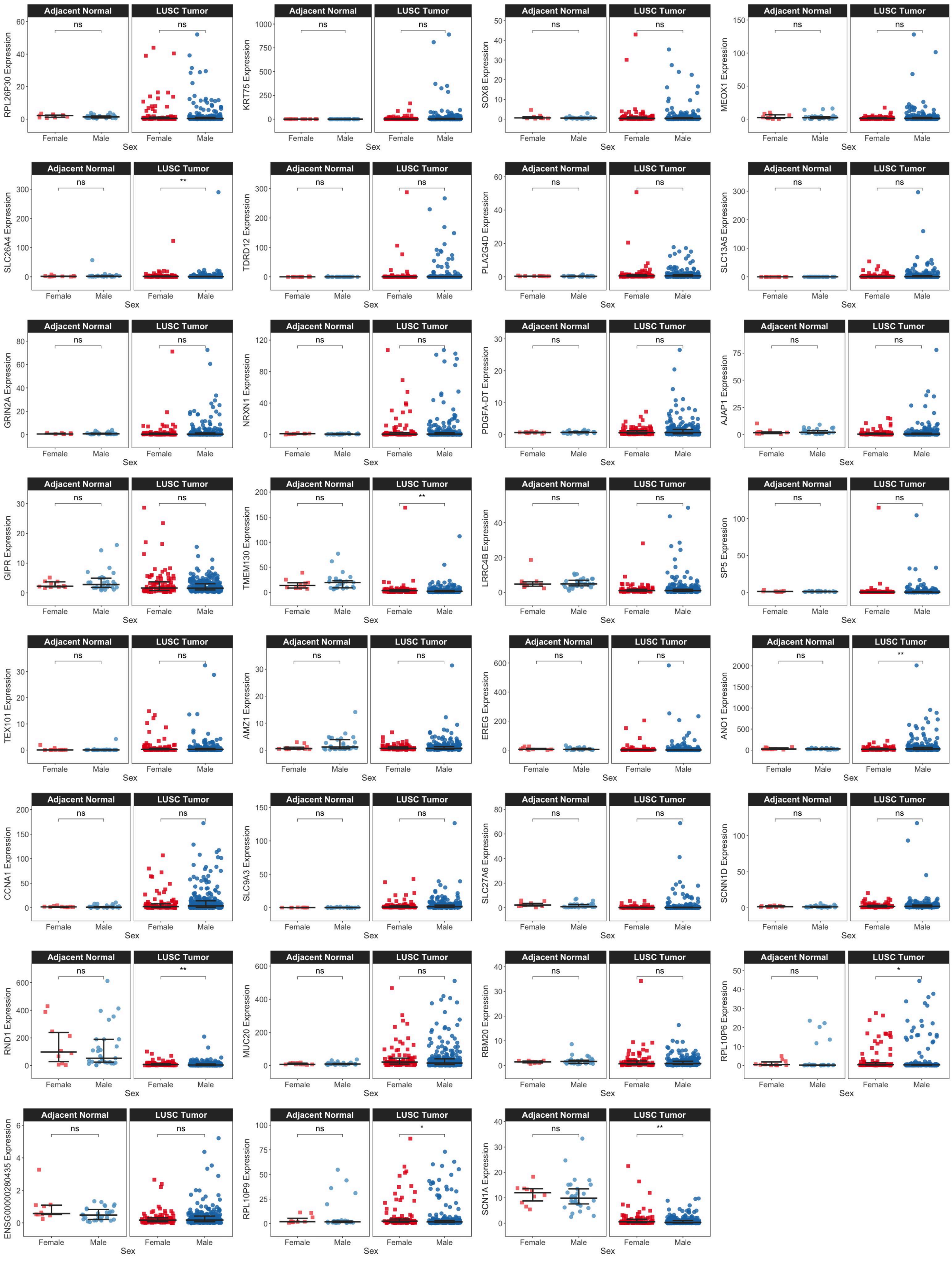


**Fig. S3**. Dot plots comparing CPM-normalized expression of male versus female tumor samples DEGs in adjacent-normal tissue and tumor samples.

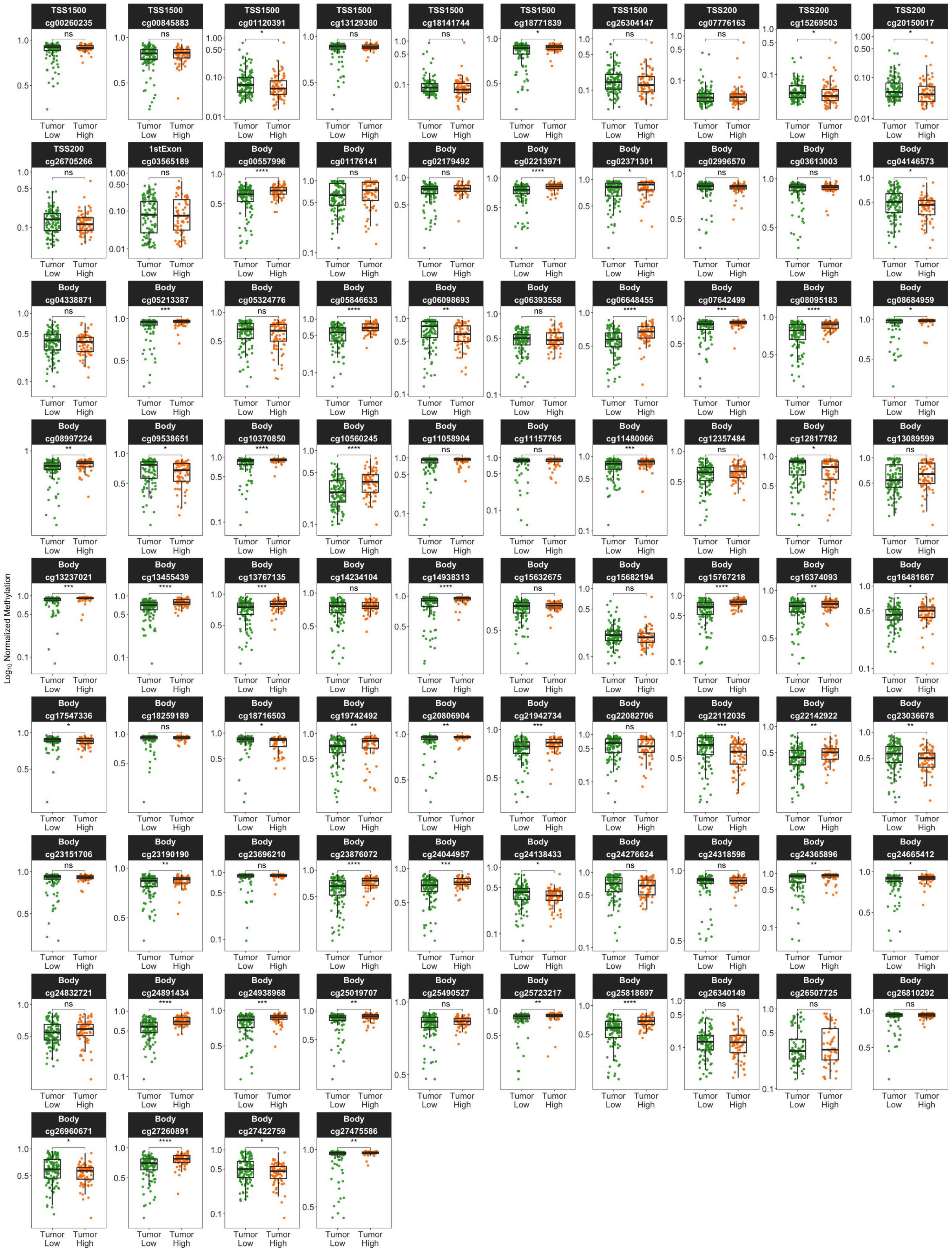


**Fig. S4**. Boxplots comparing $\log_{10}$ normalized methylation of all non-zero CpGs in the ANO1 locus (1,500 bp upstream of TSS to end of 3' UTR) for high and low ANO1-expressing male tumor samples. Dots represent individual subjects.
